# Additional Vascular Imaging in Women With Acute Cerebral Ischemia: Thresholds for Cost-Effectiveness

**DOI:** 10.64898/2026.11.19.26368544

**Authors:** Priyanka Boettger, Julian Klug, Ayush Agarwal, Omar Alhaj Omar, Martin Juenemann, Michael Buerke, Samuel Sossalla, Emmanuel Carrera

## Abstract

**Background:** Women with acute cerebral ischemia receive intracranial vascular imaging less often than men, but the value of expanding imaging is uncertain. We estimated the treatment rates and functional benefits required for additional imaging to be cost-effective.

**Methods:** We developed a decision tree and state-transition model comparing additional imaging followed by timely endovascular thrombectomy (EVT) for M1 occlusion with current care from Ontario public-payer and US Medicare perspectives. Women-specific 90-day modified Rankin Scale distributions from HERMES were projected over a lifetime. We jointly varied the proportion of additional scans leading to additional timely EVT and the treatment effect relative to HERMES. Cost-effectiveness thresholds were CAD 50,000 and USD 100,000 per quality-adjusted life-year. Program costs represented health-care resources in Ontario and additional payments under Medicare.

**Results:** At variable program costs of CAD 1,000 or USD 1,000 per additional scan, assuming the full HERMES effect and no additional fixed costs or pathway-related harm, expected cost-effectiveness required one additional timely EVT per 48 scans in Ontario and 57 under Medicare (2.07% and 1.74%, respectively). Treatment rates of 3.07% and 2.65% were required for a 90% probability of cost-effectiveness under these assumptions. At three additional thrombectomies per 100 scans and the same costs, treatment effects had to reach 76.9% and 67.3% of the HERMES effect. With the full effect, 1,000 additional scans at this treatment rate yielded approximately 5 additional functionally independent survivors at 90 days. Deterministic analyses restricted to 5 years increased the required treatment rates to 4.44% and 4.22%.

**Conclusions:** Additional vascular imaging in women could be cost-effective when a small proportion of scans leads to timely thrombectomy, provided sufficient functional benefit is retained. Prospective evaluations should assess additional treatment opportunities, functional outcomes and program costs against these conditional thresholds, which depend on projected long-term outcomes.

**What Is Known:**

- Women hospitalized with acute cerebral ischemia receive intracranial vascular imaging less often than men, although the difference is small after adjustment for clinical characteristics.
- Endovascular thrombectomy improves functional outcomes in eligible women and men, but whether extending vascular imaging to women who currently do not receive it offers good value remains unknown.

**What the Study Adds:**

- At variable program costs of CAD 1,000 or USD 1,000 per additional scan, expected cost-effectiveness required approximately one additional timely thrombectomy per 48 scans in Ontario or 57 under Medicare, assuming the full HERMES treatment effect and no additional fixed costs or pathway-related harm.
- At three additional thrombectomies per 100 scans and the same costs, the functional treatment effect had to reach approximately 77% of the HERMES effect in Ontario and 67% under Medicare.
- These findings provide conditional benchmarks for evaluating expanded imaging pathways, whose value depends on additional scans leading to timely treatment, sufficient functional benefit in the women newly reached, and the persistence of that benefit over time.

## Introduction

The benefits of endovascular thrombectomy (EVT) depend on identifying a treatable occlusion in time. Yet women with acute cerebral ischemia receive intracranial vascular imaging less often than men. In a population-based Ontario cohort of patients hospitalized within 24 hours of last seen normal, 83.6% of women and 87.8% of men underwent vascular imaging. The difference narrowed but persisted after adjustment for age, prestroke function, stroke severity, timing and hospital type (adjusted risk ratio, 0.98; 95% CI, 0.97 to 0.99).^1^ Similar differences have been reported in Alberta, Denmark and a US center.^2,3,4^ These observations raise a clinically important question: could additional imaging identify women who would otherwise miss an opportunity for effective treatment?

Randomized evidence establishes that EVT improves functional outcomes after anterior circulation large vessel occlusion, with no evidence of a difference in treatment effect by sex.^5,6^ Economic evaluations have also found thrombectomy and selected vascular imaging strategies to be cost-effective.^7,8,9,10,11,12,13^ These findings do not establish the value of extending imaging to women who currently do not receive it. That value depends on two linked quantities: how often additional imaging leads to additional timely EVT, and how much the women treated benefit. Women reached through an expanded pathway may differ from trial participants in prestroke function, comorbidity and time to treatment, making direct transfer of trial benefit uncertain.

Neither quantity is established for the 1,307 women in the Ontario cohort who received no vascular imaging.^1^ The observed imaging difference cannot distinguish missed treatment opportunities from appropriate clinical selection, and occlusion prevalence among women already imaged cannot resolve this uncertainty. Previous economic work on sex differences in stroke care has examined improved prehospital recognition.^14^ The in-hospital decision requires evidence on when expanding access to imaging would produce sufficient health gains to justify the additional resources.

We therefore estimated the minimum rates of additional timely EVT and the functional treatment effects required for additional vascular imaging in women to be cost-effective. We evaluated these requirements jointly from Ontario public-payer and US Medicare perspectives across program costs, and examined their sensitivity to the duration and assumptions of projected long-term outcomes. The aim was to define measurable conditions for a worthwhile imaging pathway that could guide its prospective evaluation.

## Methods

### Study Design and Population

We conducted a conditional cost-effectiveness and threshold analysis^15^ of additional intracranial vascular imaging for women with acute cerebral ischemia, compared with current care, from the Ontario public-payer and the US Medicare perspective. Because the additional treatment rate and the treatment effect in women who currently do not receive imaging are unknown, we estimated the combinations of these quantities required for cost-effectiveness. Reporting followed CHEERS 2022.^16^ The protocol was finalized on 18 October 2026 before results were generated; six amendments made after results were known are documented, with their consequences, in Supplemental Methods 5. The study used published aggregate data and did not require research ethics review.

The source cohort comprised 7,967 women hospitalized in Ontario with cerebral ischemia within 24 hours of last seen normal, including 1,307 who received no intracranial vascular imaging.^1^ The modeled pathway offered computed tomography angiography to women with suspected ischemic stroke in whom hemorrhage had been excluded and no angiography had been ordered, unless a documented exclusion applied, followed by rapid interpretation and referral for EVT (Figure 1). Women entered the model at age 78 years, the median age of Ontario women with large vessel occlusion. The primary analysis credited treatment benefit for M1 occlusion, while the HERMES effect pools anterior circulation occlusion locations; basilar occlusion was examined in a scenario and no benefit was assigned to EVT for medium vessel occlusion.^17^ Clinical inputs and their applicability are summarized in Table 1; occlusion frequencies, population definitions and exclusions are in Supplemental Methods 1.

**Figure 1.**
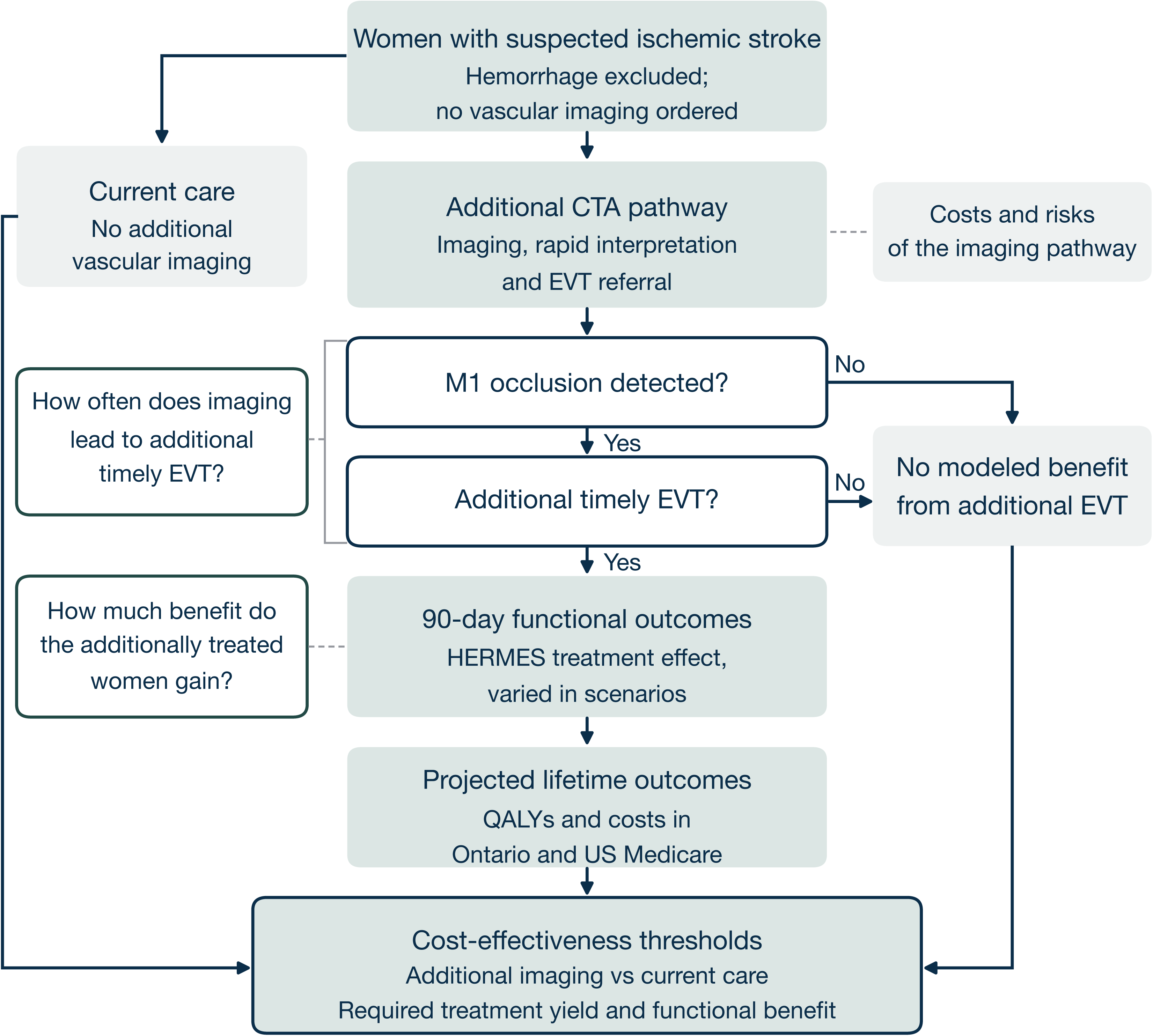
Clinical pathway, the two unknown quantities and the comparison. Women with suspected ischemic stroke in whom noncontrast computed tomography has excluded hemorrhage and no vascular imaging has been ordered either continue under current care or enter the additional computed tomography angiography (CTA) pathway; both arms meet in the same comparison. Solid arrows are modeled paths, dashed grey lines are explanatory notes. Treatment yield denotes additional timely thrombectomies per additional scan in women with confirmed cerebral ischemia; a yield of 3% represents three additional timely thrombectomies per 100 such scans, compared with current care, and is an assumed scenario, not an observed pathway yield. It therefore spans both decisions drawn here, whether an M1 occlusion is detected and whether that leads to additional timely endovascular thrombectomy (EVT), which is why the note is tied to both. The second scenario axis is the share of the women-specific HERMES treatment effect retained by the additionally treated women. The primary model credits additional EVT-related benefit only for M1 occlusion, the M1 segment of the middle cerebral artery; benefits from other management changes after imaging are not quantified, and basilar occlusion enters only in a scenario. Program costs and any modeled pathway-related harm apply across all additional scans, including those followed by EVT, and the program cost also covers imaging and assessments in women whose final diagnosis is not cerebral ischemia; the primary analysis assumes no pathway-related harm. Ninety-day modified Rankin Scale (mRS) distributions come from the women in HERMES (randomized, EVT versus control) and are projected to age 110 with Canadian-adjusted Oxford Vascular Study transitions, female life-table mortality plus mRS-specific excess mortality, utilities and costs by disability state, valued as Ontario public-payer resources or as Medicare payments (Supplemental Methods 2 to 4). The detailed model diagram, with the symbols of the model drawn in, is Figure S8. QALY indicates quality-adjusted life-year.

**Table 1.**
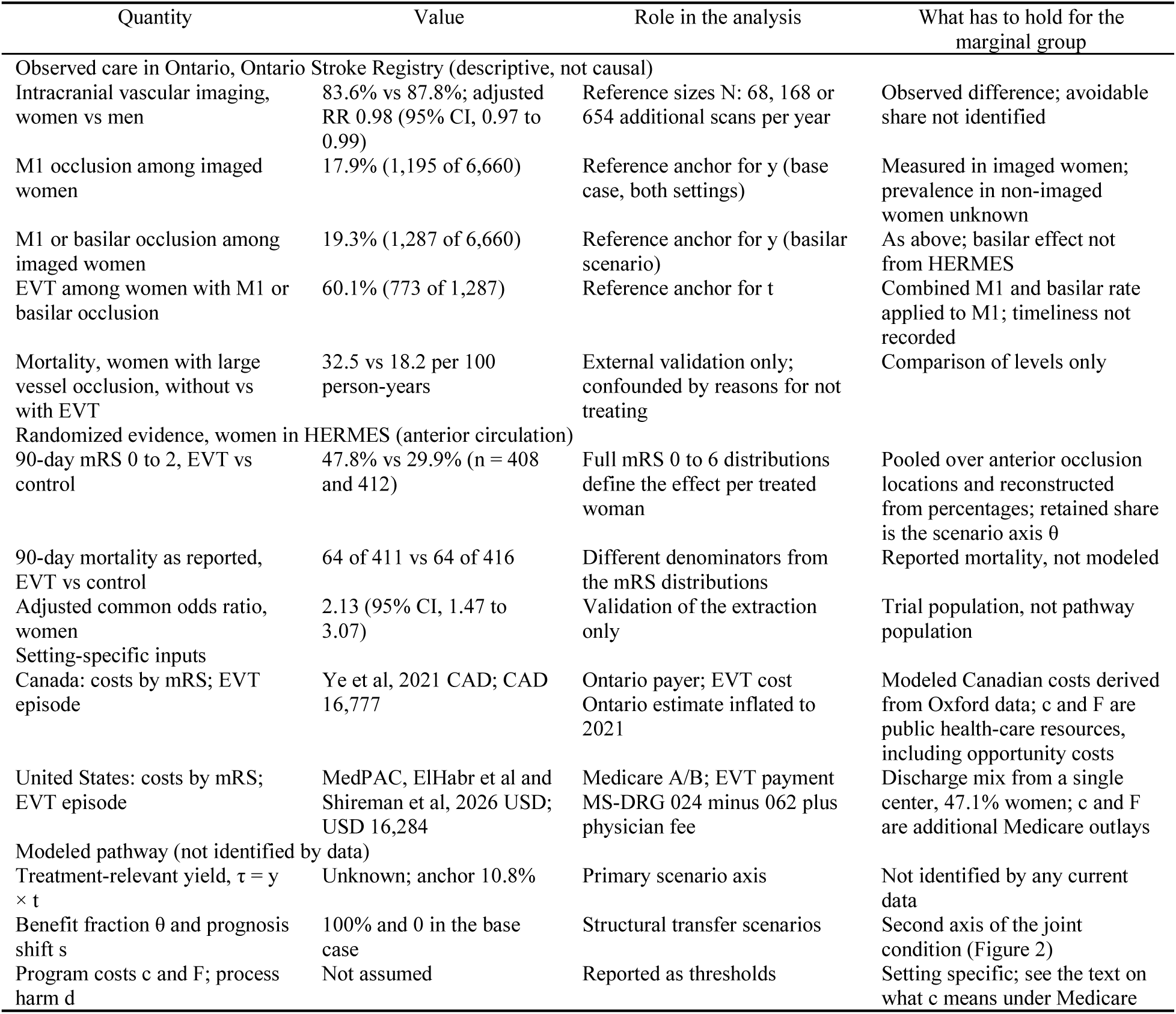
Three kinds of evidence and how each enters the analysis. Observed values are from the Ontario Stroke Registry cohort of patients hospitalized with cerebral ischemia within 24 hours of last seen normal and serve as reference values in both settings; randomized values are from women in the HERMES individual patient data meta-analysis. Sources for every parameter are in the Supplemental Material. CAD indicates Canadian dollars; EVT, endovascular thrombectomy; mRS, modified Rankin Scale; RR, risk ratio; and USD, US dollars. The last column states what has to hold for each value to apply to the women a pathway would additionally reach; women specific is not the same as specific to that marginal group.

| Quantity | Value | Role in the analysis | What has to hold for the marginal group |
|---|---|---|---|
| Observed care in Ontario, Ontario Stroke Registry (descriptive, not causal) |  |  |  |
| Intracranial vascular imaging, women vs men | 83.6% vs 87.8%; adjusted RR 0.98 (95% CI, 0.97 to 0.99) | Reference sizes N: 68, 168 or 654 additional scans per year | Observed difference; avoidable share not identified |
| M1 occlusion among imaged women | 17.9% (1,195 of 6,660) | Reference anchor for y (base case, both settings) | Measured in imaged women; prevalence in non-imaged women unknown |
| M1 or basilar occlusion among imaged women | 19.3% (1,287 of 6,660) | Reference anchor for y (basilar scenario) | As above; basilar effect not from HERMES |
| EVT among women with M1 or basilar occlusion | 60.1% (773 of 1,287) | Reference anchor for t | Combined M1 and basilar rate applied to M1; timeliness not recorded |
| Mortality, women with large vessel occlusion, without vs with EVT | 32.5 vs 18.2 per 100 person-years | External validation only; confounded by reasons for not treating | Comparison of levels only |
| Randomized evidence, women in HERMES (anterior circulation) |  |  |  |
| 90-day mRS 0 to 2, EVT vs control | 47.8% vs 29.9% (n = 408 and 412) | Full mRS 0 to 6 distributions define the effect per treated woman | Pooled over anterior occlusion locations and reconstructed from percentages; retained share is the scenario axis θ |
| 90-day mortality as reported, EVT vs control | 64 of 411 vs 64 of 416 | Different denominators from the mRS distributions | Reported mortality, not modeled |
| Adjusted common odds ratio, women | 2.13 (95% CI, 1.47 to 3.07) | Validation of the extraction only | Trial population, not pathway population |
| Setting-specific inputs |  |  |  |
| Canada: costs by mRS; EVT episode | Ye et al, 2021 CAD; CAD 16,777 | Ontario payer; EVT cost Ontario estimate inflated to 2021 | Modeled Canadian costs derived from Oxford data; c and F are public health-care resources, including opportunity costs |
| United States: costs by mRS; EVT episode | MedPAC, ElHabr et al and Shireman et al, 2026 USD; USD 16,284 | Medicare A/B; EVT payment MS-DRG 024 minus 062 plus physician fee | Discharge mix from a single center, 47.1% women; c and F are additional Medicare outlays |
| Modeled pathway (not identified by data) |  |  |  |
| Treatment-relevant yield, τ = y × t | Unknown; anchor 10.8% | Primary scenario axis | Not identified by any current data |
| Benefit fraction θ and prognosis shift s | 100% and 0 in the base case | Structural transfer scenarios | Second axis of the joint condition (Figure 2) |
| Program costs c and F; process harm d | Not assumed | Reported as thresholds | Setting specific; see the text on what c means under Medicare |

All pathway quantities were expressed per additional scan in a woman with a final diagnosis of cerebral ischemia. Costs of assessments and scans in women with other final diagnoses were included in the program cost on this denominator; associated health losses count as pathway-related harm.

### Clinical Outcomes and Treatment Benefit

A decision tree linked additional imaging to the detection of an M1 occlusion and to timely EVT (Figure 1). The treatment-relevant yield was the proportion of additional scans leading to an additional timely thrombectomy; women without additional timely EVT were assumed to have the same outcomes as under current care, apart from any pathway-related harm.

Treatment outcomes were derived from the women in the EVT and control groups of HERMES with 90-day modified Rankin Scale (mRS) scores (408 and 412 women), reconstructed from the published percentages.^6^ A state-transition model projected these distributions through mRS states 0 to 5 and death to age 110, using Canadian-adjusted Oxford Vascular Study transitions,^18^ female life-table mortality with an mRS-specific excess hazard,^19,20^ utilities^21^ and costs by disability state. Outcomes beyond 90 days are therefore modeled projections (Supplemental Methods 2; Tables S1 to S4).

We varied the treatment-relevant yield and the functional treatment effect relative to HERMES jointly. A reduced effect was modeled by proportionally reducing the difference between the EVT and the control mRS distributions while retaining the full EVT episode cost; this fraction describes the magnitude of the treatment effect, not the proportion of women who benefit. Both quantities were scenario inputs because their values among the women reached by additional imaging are unknown; registry-derived values served only as references (Supplemental Methods 1). Older age (86 years) and a worse baseline prognosis (25% or 50% of women in both groups one mRS category worse) were separate scenarios.

### Costs and Cost-Effectiveness Thresholds

Ontario costs represent public health-care resources, including resources absorbed within existing budgets, in 2021 Canadian dollars, with costs by disability state from Ye et al^18^ and an incremental EVT episode cost of CAD 16,777.^7,11^ Costs and health outcomes were discounted at 1.5% annually, with a willingness to pay of CAD 50,000 per quality-adjusted life-year (QALY).^22^ The US analysis represents Medicare Parts A and B payments in 2026 US dollars, with 3% annual discounting and a threshold of USD 100,000 per QALY,^23,24^ and an incremental EVT payment of USD 16,284.^8,25,26,27,28,29^ Program costs under Medicare are additional outlays, whereas in Ontario they value health-care resources. Payment derivations, alternative cost assumptions and the separate hospital perspective are in Supplemental Methods 4 and Tables S4 to S8.

We calculated the incremental net monetary benefit as the value of the additional QALYs at the cost-effectiveness threshold of each setting minus the additional costs: per additionally treated woman λΔQ − ΔC, and per additionally imaged woman the yield times that amount, less the program cost and any pathway-related harm. We identified the minimum additional timely EVT yield and the minimum retained treatment effect required for a nonnegative expected net monetary benefit, together with the thresholds associated with a 90% probability of cost-effectiveness, and varied the two quantities jointly (Table 2; Figure 2). The primary thresholds assume no additional fixed program costs and no pathway-related harm, both examined separately; equations and derivations are in Supplemental Methods 2.

**Figure 2.**
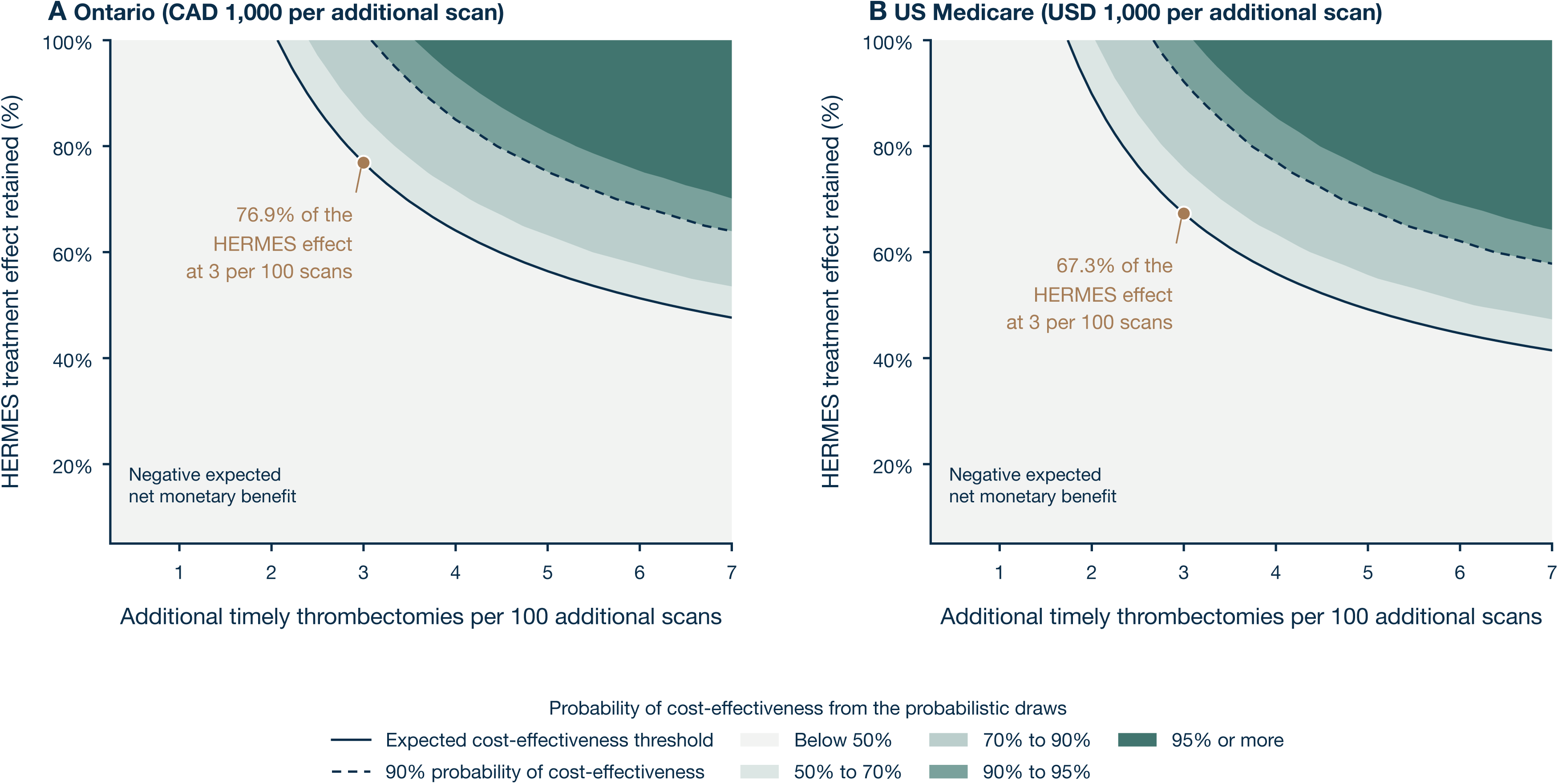
Yield and retained treatment effect as one joint condition, at 1,000 per additional scan. A, Ontario public payer (2021 Canadian dollars); B, US Medicare (2026 US dollars). The horizontal axis is the additional timely thrombectomy yield, that is additional timely thrombectomies per additional scan in a woman with confirmed cerebral ischemia, shown as additional timely thrombectomies per 100 additional scans; the vertical axis is the share of the women-specific HERMES functional benefit retained by the additionally treated women. Treatment yield denotes additional timely thrombectomies enabled by the pathway per 100 additional scans in women with confirmed cerebral ischemia; it is a scenario assumption, not an observed rate, and so is the retained treatment effect. The solid line is the expected cost-effectiveness threshold, where the mean net monetary benefit is zero; the dashed line is the yield and retained effect at which the pathway is cost-effective in 90% of the probabilistic draws. The graded shading is the probability of cost-effectiveness computed from the draws at each combination, in five bands (below 50%, 50% to 70%, 70% to 90%, 90% to 95%, 95% or more); it is not derived from the expected threshold, which need not coincide with a probability of 50%. The marked point shows the minimum retained HERMES treatment effect required for expected cost-effectiveness at three additional timely thrombectomies per 100 scans, 76.88% and 67.28%. Fixed program costs and pathway-related harm are zero in both panels; either one raises the yield required, and the room for fixed costs is in Table 2. These are thresholds of this model at a fixed program cost, not estimates of what a pathway would achieve. EVT indicates endovascular thrombectomy.

**Table 2.**
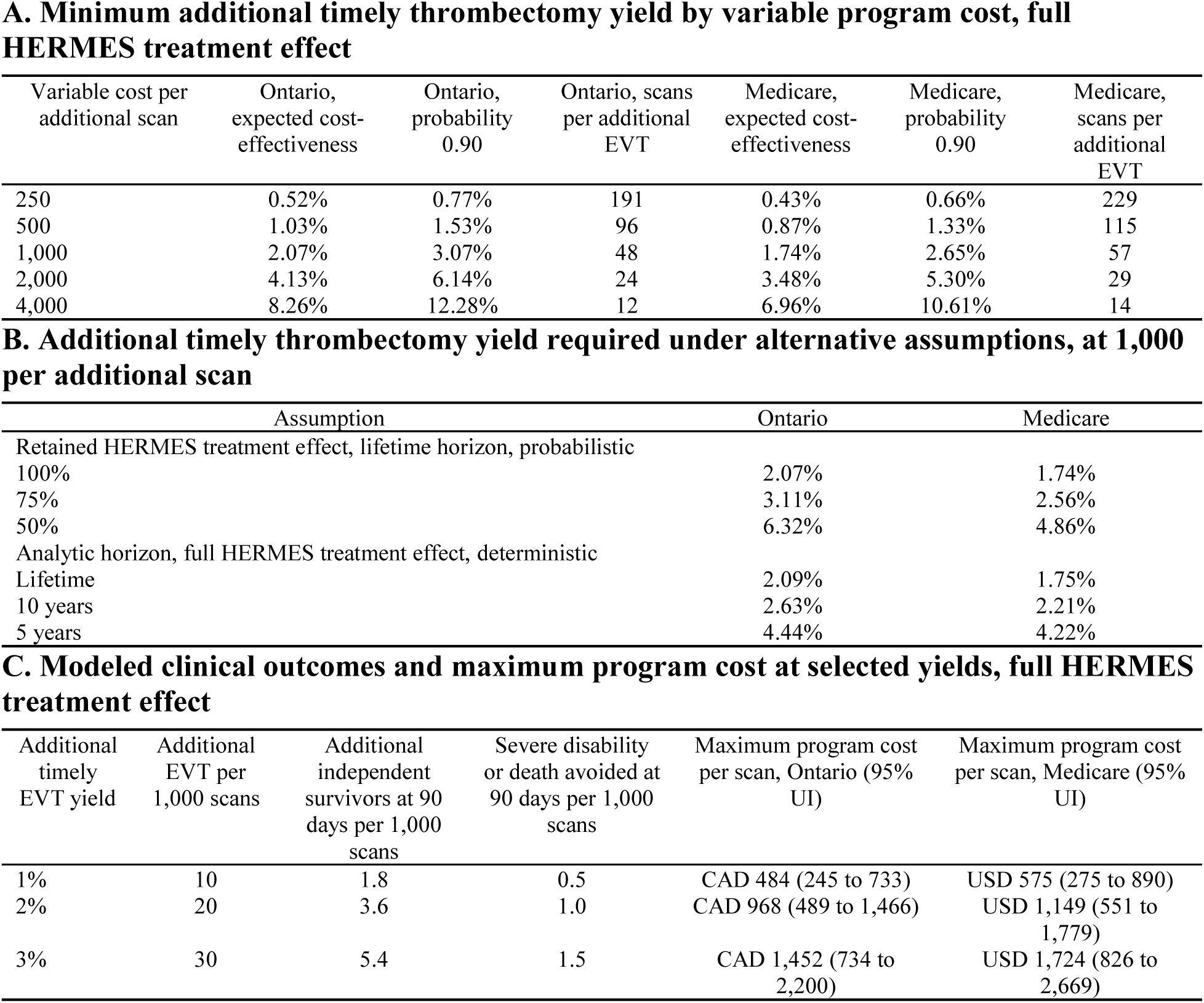
Treatment requirements, clinical outcomes and program cost limits for additional vascular imaging in women with acute cerebral ischemia. Costs are in 2021 Canadian dollars for Ontario and 2026 US dollars for Medicare. Yield is additional timely thrombectomies per additional scan in a woman with confirmed cerebral ischemia; the variable cost per additional scan includes all incremental costs of the rule (scans and assessments in women with other final diagnoses, staff time, reading, transfer coordination). Expected cost-effectiveness is the yield at which the mean net monetary benefit across 16,000 and 23,000 draws reaches zero; probability 0.90 is the yield at which the pathway is cost-effective in 90% of draws; scans per additional EVT is the reciprocal of the expected break-even yield. Panels A to C assume no pathway-related harm, and Panels A and B no additional fixed program cost. In Panel B the retained treatment effect is the size of the functional benefit relative to HERMES, not the proportion of women who benefit, and its rows are probabilistic means; the full curve is in Figure S5 and Table S12. The horizon rows of Panel B are deterministic and limit how long the modeled treatment difference is counted, so they carry their own lifetime reference, which differs slightly from the probabilistic lifetime value above (Figure 4, Table S11). In Panel C, independence is mRS 0 to 2 and severe disability or death is mRS 5 to 6; these clinical quantities are from the Ontario draws and differ from the Medicare values only beyond the precision reported here. The maximum program cost is the highest total cost per additional scan, variable and fixed together, at which the pathway remains cost-effective (probabilistic mean, 95% uncertainty interval). The zero-cost limiting case, what an additional scan costs under bundled US hospital payment, and the process-harm thresholds are in Table S21. All values are thresholds of this model, not estimates of what a pathway would achieve. Ontario: CAD 50,000 per quality-adjusted life-year, 1.5% discounting; Medicare: USD 100,000, 3%; lifetime horizon. EVT indicates endovascular thrombectomy; mRS, modified Rankin Scale; and UI, uncertainty interval.

| **A. Minimum additional timely thrombectomy yield by variable program cost, full HERMES treatment effect** |  |  |  |  |  |  |
|---|---|---|---|---|---|---|
| Variable cost per additional scan | Ontario, expected cost-effectiveness | Ontario, probability 0.90 | Ontario, scans per additional EVT | Medicare, expected cost-effectiveness | Medicare, probability 0.90 | Medicare, scans per additional EVT |
| 250 | 0.52% | 0.77% | 191 | 0.43% | 0.66% | 229 |
| 500 | 1.03% | 1.53% | 96 | 0.87% | 1.33% | 115 |
| 1,000 | 2.07% | 3.07% | 48 | 1.74% | 2.65% | 57 |
| 2,000 | 4.13% | 6.14% | 24 | 3.48% | 5.30% | 29 |
| 4,000 | 8.26% | 12.28% | 12 | 6.96% | 10.61% | 14 |
| **B. Additional timely thrombectomy yield required under alternative assumptions, at 1,000 per additional scan** |  |  |  |  |  |  |
| Assumption | Ontario | Medicare |  |  |  |  |
| Retained HERMES treatment effect, lifetime horizon, probabilistic |  |  |  |  |  |  |
| 100% | 2.07% | 1.74% |  |  |  |  |
| 75% | 3.11% | 2.56% |  |  |  |  |
| 50% | 6.32% | 4.86% |  |  |  |  |
| Analytic horizon, full HERMES treatment effect, deterministic |  |  |  |  |  |  |
| Lifetime | 2.09% | 1.75% |  |  |  |  |
| 10 years | 2.63% | 2.21% |  |  |  |  |
| 5 years | 4.44% | 4.22% |  |  |  |  |
| **C. Modeled clinical outcomes and maximum program cost at selected yields, full HERMES treatment effect** |  |  |  |  |  |  |
| Additional timely EVT yield | Additional EVT per 1,000 scans | Additional independent survivors at 90 days per 1,000 scans | Severe disability or death avoided at 90 days per 1,000 scans | Maximum program cost per scan, Ontario (95% UI) | Maximum program cost per scan, Medicare (95% UI) |  |
| 1% | 10 | 1.8 | 0.5 | CAD 484 (245 to 733) | USD 575 (275 to 890) |  |
| 2% | 20 | 3.6 | 1.0 | CAD 968 (489 to 1,466) | USD 1,149 (551 to 1,779) |  |
| 3% | 30 | 5.4 | 1.5 | CAD 1,452 (734 to 2,200) | USD 1,724 (826 to 2,669) |  |

### Scenarios, Program Scale and Budget Impact

Because the projection carries most of the modeled gain, we truncated the analytic horizon to 10, 5, 2 and 1 years and varied the extrapolation of mortality and functional state. Further scenarios varied age, prognosis, treatment effect, discounting, willingness to pay and costs (Tables S7, S8, S10 and S11). For Ontario we considered three program sizes, up to all 654 currently non-imaged women per year, and calculated the maximum fixed annual cost consistent with cost-effectiveness (Supplemental Methods 2; Table S17). Budget impact was calculated for a single annual cohort over 1 and 5 years and, for a program operating over several years, as the program costs of that year plus the downstream costs of all cohorts enrolled up to that year (Tables S15, S16 and S18).

### Uncertainty and Model Assessment

Probabilistic analyses propagated uncertainty in the clinical outcomes, transitions, utilities, mortality and costs, with draws added until prespecified precision was reached; distributions, dependencies, convergence criteria and the effective sample size of the transition matrices are in Supplemental Methods 3 and Tables S1, S9 and S13. Yield, the transfer of the treatment effect, program size, program costs and pathway-related harm were evaluated as scenarios, not as probability distributions: the probabilistic analysis describes parameter uncertainty and gives no probability that a pathway attains a given yield or retains a given share of the benefit.

Model checks covered internal consistency, limiting cases, the threshold calculations and external plausibility; they and the implementation of the base R model (4.6.0) are in Supplemental Methods 6 and Table S19. Generative artificial intelligence assistants were used during this work: Microsoft Copilot (Microsoft) to improve language, grammar and readability, and Claude (Anthropic) to review analysis code written by the authors; the authors verified every input against its source and made all scientific decisions. Results are probabilistic means with 95% uncertainty intervals (UIs) unless identified as deterministic. The authors had full access to all data and take responsibility for their integrity and analysis.

## Results

### Treatment Requirements for Cost-Effectiveness

At variable program costs of CAD 1,000 in Ontario and USD 1,000 under Medicare per additional scan, expected cost-effectiveness required about one additional timely thrombectomy per 48 scans in Ontario and per 57 under Medicare, corresponding to yields of 2.07% and 1.74%. These thresholds assume the full HERMES treatment effect and no additional fixed costs or pathway-related harm. Yields of 3.07% and 2.65% were required for a 90% probability of cost-effectiveness (Table 2; Figure 3).

**Figure 3.**
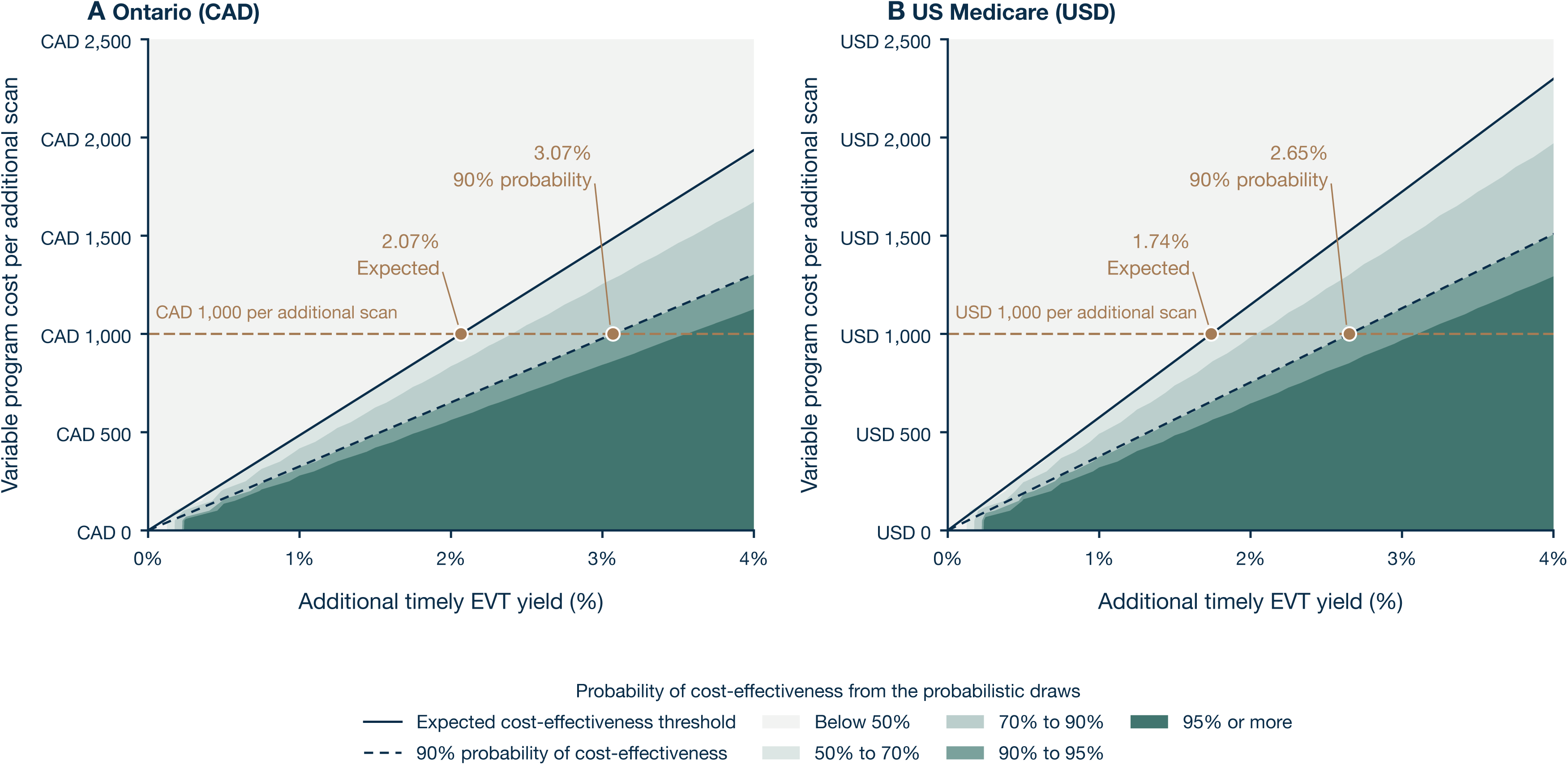
Variable program cost and the yield required, in the decision-relevant range. A, Ontario public payer (2021 Canadian dollars, 1.5% discounting, CAD 50,000 per quality-adjusted life-year); B, US Medicare (2026 US dollars, 3% discounting, USD 100,000). Combinations of additional timely thrombectomy yield and variable program cost per additional scan at which the pathway is cost-effective: the solid line is the expected cost-effectiveness threshold and the dashed line the threshold at which the pathway is cost-effective in 90% of the probabilistic draws, with the same five graded probability bands as Figure 2, computed from the 16,000 and 23,000 draws of the two settings. The horizontal dashed line marks 1,000 per additional scan, and the marked points are the expected and 90%-probability yields at that cost. Fixed program costs and pathway-related harm are zero throughout and either one raises the yield required. Women aged 78 years. The full range of yields, with the derived M1 reference value in currently imaged women, is Figure S9. EVT indicates endovascular thrombectomy.

The required yield rose as the functional benefit declined. At three additional thrombectomies per 100 scans, cost-effectiveness required 76.88% and 67.28% of the HERMES treatment effect, that is of the size of the additional functional gain over no thrombectomy. Halving the treatment effect increased the required yields to 6.32% and 4.86% (Table 2; Figure 2). Retained benefit fractions below which no yield of 100% or less suffices are in Table S12.

### Projected Clinical Outcomes

At a yield of 3%, that is three additional thrombectomies per 100 additional scans, the model projected 30 additional timely thrombectomies, about five additional women alive and functionally independent (mRS 0 to 2) at 90 days and 1.5 fewer women dead or severely disabled per 1,000 additional scans in either setting (Table 2; Figure S6). Per additionally treated woman, EVT gained 0.97 (95% UI, 0.58 to 1.39) QALYs in Ontario and 0.83 (95% UI, 0.49 to 1.17) under Medicare, at incremental lifetime costs of CAD 331 (95% UI, −CAD 10,943 to CAD 10,718) and USD 25,155 (95% UI, USD 16,856 to USD 34,560) (Tables S6 and S9).

### Sensitivity to Long-Term Outcomes and Patient Characteristics

About 89% to 90% of the projected lifetime QALY gain accrued after the first year. In deterministic analyses at CAD 1,000 and USD 1,000 per scan, restricting the horizon to 10 years increased the required yield to 2.63% and 2.21% and a 5-year horizon to 4.44% and 4.22% (Table 2; Figure 4); over 1 year the net monetary benefit per treated woman was negative in both settings. Alternative mortality and functional-transition assumptions moved the thresholds in both directions (Table S11). Moving 50% of women in both groups one mRS category worse increased the expected thresholds to 2.37% in Ontario and 1.92% under Medicare, and combining that shift with age 86 years and half the HERMES effect to 17.35% and 12.90% (Tables S8 and S10).

**Figure 4.**
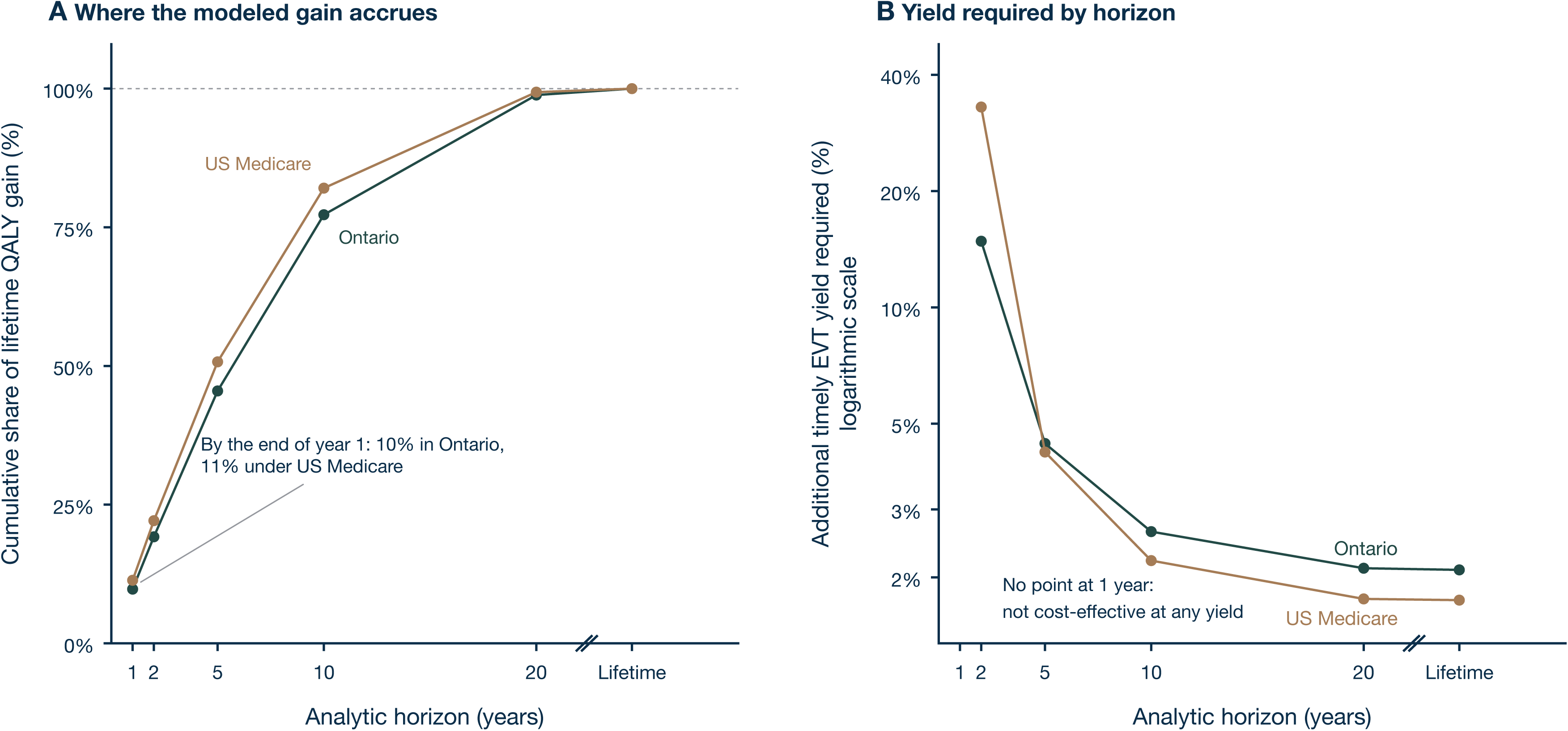
Time horizon and the value of additional imaging, deterministic at 1,000 per additional scan in the currency of each setting. A, Cumulative share of the modeled lifetime quality-adjusted life-year (QALY) gain per treated woman that has accrued by the end of each analytic horizon, in Ontario and under US Medicare; the dashed line marks the lifetime total. B, Additional timely thrombectomy yield at which the pathway reaches the cost-effectiveness threshold, by horizon, on a logarithmic vertical scale. The lifetime horizon is set off from the numeric years by an axis break in both panels. In B no point is drawn at 1 year: within 1 year the net monetary benefit per treated woman is negative in both settings, so no yield is sufficient. Both panels are deterministic, so the lifetime values (2.09% in Ontario and 1.75% under Medicare in B) differ slightly from the probabilistic means reported in the text and in Table 2. The horizon limits the duration of the modeled treatment difference; values are in Table S11. EVT indicates endovascular thrombectomy.

### Program Costs and Budget Impact

For an Ontario program adding 68 scans per year at CAD 1,000 per scan, the maximum fixed annual cost consistent with cost-effectiveness was CAD 30,700 at a 3% yield; at a 2% yield the variable costs already exceeded the modeled value by about CAD 2,200 (Table S17). In a program that admits women every year, the additional payer cost per 1,000 additional scans at a 3% yield moved from CAD 1,145,600 in the first calendar year to CAD 978,100 in the fifth in Ontario, where the later years of earlier cohorts bring savings, and from USD 1,435,000 to USD 1,555,000 under Medicare, where they add payments (Table S18; Figure S7). Downstream costs of a single annual cohort are in Table S15, and the separate hospital revenue analysis in Supplemental Methods 4. The two settings differ in currency, willingness to pay and cost perspective, so their thresholds are not a comparison of health-system efficiency.

The probabilistic analyses met the precision criteria and every recorded model check passed. Modeled survival and mortality were of the same order as those observed in Ontario women with large vessel occlusion;^1^ such comparisons support the plausibility of the projection but do not validate outcomes in women reached by an additional-imaging pathway (Tables S9 and S19).

## Discussion

Additional vascular imaging in women with acute cerebral ischemia could be cost-effective at low rates of additional timely thrombectomy, provided the women newly treated achieve sufficient functional benefit. At variable program costs of CAD 1,000 in Ontario and USD 1,000 under Medicare per additional scan, three additional thrombectomies per 100 scans required 76.9% and 67.3% of the HERMES treatment effect, assuming no additional fixed costs or pathway-related harm. These findings define measurable treatment and outcome requirements for evaluating expanded imaging pathways. The economic value of expanded imaging depends on the additional effective treatment it enables and the functional benefit achieved in the women newly treated.

Previous economic evaluations have supported vascular imaging in defined stroke populations. Wu et al examined computed tomography angiography in patients with minor stroke and reported a threshold for large vessel occlusion prevalence, whereas Martinez-Gutierrez et al compared initial imaging strategies at comprehensive stroke centers.^12,13^ Both evaluated imaging within specified populations and treatment pathways. Economic evaluations of stroke quality measures have similarly shown that the potential value of improving care varies across processes.^30^ Gadsden et al examined sex differences earlier in the pathway, modeling improved prehospital recognition and access to thrombolysis among Australian women younger than 70 years.^14^ Our analysis addresses the women an additional in-hospital imaging pathway would reach, whose likelihood of receiving timely EVT and capacity to benefit may differ from those of women already imaged or enrolled in thrombectomy trials. Varying these two quantities jointly makes their implications for cost-effectiveness explicit.

The projected clinical outcomes illustrate the scale of benefit associated with these requirements. At three additional thrombectomies per 100 scans and the full HERMES effect, the model projected about five additional women alive and functionally independent at 90 days per 1,000 additional scans. About 89% to 90% of the lifetime QALY gain accrued after the first year, through the time subsequently spent in different functional states and the modeled difference in survival (Figure 4). This dependence on longer-term outcomes is consistent with previous thrombectomy evaluations, in which cost-effectiveness differed substantially between trial and lifetime horizons.^10^ Restricting the horizon to 5 years more than doubled the required treatment rate in both settings, to 4.44% and 4.22%. A shorter analytic horizon limits the outcomes that are counted; it does not establish when the treatment difference ends.

The observed sex difference provides a rationale for examining access to imaging, but does not establish how much of that difference represents missed treatment opportunities. In the Ontario cohort the imaging difference was small after adjustment, and EVT use was similar among women and men with a detected large vessel occlusion.^1^ Similar treatment rates after occlusion detection do not resolve potential differences earlier in the diagnostic pathway, and the documented reasons for withholding thrombectomy do not explain why vascular imaging was not performed. Prospective evaluation should therefore document clinical eligibility and the reasons for withholding imaging alongside additional timely treatment and patient outcomes. The budget consequences also matter for implementation. Under Medicare the projected health gains justified the additional expenditure at the selected willingness to pay, although lifetime payments increased. In Ontario the allowance for fixed implementation costs was limited at the lower treatment rates examined. A pathway may therefore meet a cost-effectiveness threshold while requiring additional funding or leaving little scope for implementation expenditure, so decisions about adoption should weigh the expected health gains against the resources required for imaging, interpretation, referral and treatment.

### Implications for Prospective Evaluation

Prospective studies should measure additional imaging, additional timely EVT, functional outcomes and the full costs of implementation, including the assessment of patients whose final diagnosis is not cerebral ischemia. A concurrent comparison is needed to distinguish treatment enabled by the pathway from treatment that would have occurred under usual care.

A randomized implementation, in a parallel-group or stepped-wedge design, could estimate the overall effect of the pathway,^31^ while analyses of prestroke function, stroke severity, occlusion location and time to treatment could inform the applicability of HERMES,^32^ although outcomes among additionally treated women alone cannot identify the causal treatment effect because this group is selected by the pathway. Follow-up beyond 90 days would inform the persistence of functional differences and subsequent resource use, and would allow the thresholds derived here to be compared with achievable treatment rates, outcomes and costs.

### Limitations

The additional timely treatment rate and treatment effect in women reached by expanded imaging are unknown. The analysis defines cost-effectiveness conditions, but not the probability that a pathway will meet them. Long-term outcomes were extrapolated from 90-day functional distributions; sensitivity analyses cannot validate the projected duration of benefit.

HERMES distributions were reconstructed from published percentages across anterior circulation occlusions, whereas the primary model credited only benefits from additional EVT for M1 occlusion. Benefits from other management changes following imaging were not quantified. Trial participants may differ from newly reached women, and proportional effect attenuation cannot capture all differences in treatment response. The registry lacked imaging timing, excluded smaller hospitals, covered confirmed cerebral ischemia rather than all suspected strokes, and predated recent expansion of the evidence supporting EVT, potentially limiting contemporary applicability.^1^

Costs combined published sources, including a single-center discharge distribution for US post-acute care. No implemented pathway supplied program costs, and pathway-related harm was assessed through thresholds. Inputs were identified through targeted searches rather than systematic evidence synthesis; relevant studies may have been missed, and alternative source selection could affect the estimated thresholds (Supplemental Methods 7).

### Conclusions

Additional vascular imaging in women with acute cerebral ischemia could be cost-effective even when few additional scans lead to timely thrombectomy, but the required treatment rate rises substantially as functional benefit declines. These joint thresholds provide benchmarks for prospective evaluation, conditional on program costs and projected long-term outcomes. Whether expanded imaging can meet these requirements in women currently not imaged remains to be established.

## Acknowledgments

P.Bo. gratefully acknowledges the Department of Health Policy, London School of Economics and Political Science, and the support of a European Heart Academy Scholarship. Generative artificial intelligence assistants were used during this work: Microsoft Copilot (Microsoft) to improve language, grammar and readability, and Claude (Anthropic) to review analysis code written by the authors. The authors reviewed and verified all AI-assisted revisions. No image is created or enhanced with AI-tools. All scientific decisions were made by the authors, who verified every input against its source and take full responsibility for the content.

## Declarations

### Author Contributions

Dr Boettger conceived the study, specified the decision problem, built the model and the parameter register, wrote the analysis code, carried out the analyses and drafted the manuscript. Drs Klug, Agarwal, Alhaj Omar, Juenemann, Buerke, Sossalla and Carrera contributed to the clinical and pathway assumptions and to the interpretation of the results, and revised the manuscript critically. Dr Carrera supervised the work. All authors approved the submitted version and accept responsibility for its content.

### Data Availability

This analysis used published aggregate data only; no individual patient data were obtained, and the registry and trial values it reuses are available in the cited publications. The model code, the parameter register with every input, its range, distribution and source, the raw input files that may be redistributed and all output tables, including the probabilistic draws, are provided with this article so that every reported number can be regenerated from one run.

### Supplemental Material

Supplemental Methods 1 to 7; Figures S1 to S9; Tables S1 to S22, including the CHEERS 2022 checklist; model code, parameter register and output tables.

## Nonstandard Abbreviations and Acronyms

CTA: computed tomography angiography
EVT: endovascular thrombectomy
HERMES: Highly Effective Reperfusion Evaluated in Multiple Endovascular Stroke Trials
mRS: modified Rankin Scale
QALY: quality-adjusted life-year
UI: uncertainty interval

## References

1. Siddharthan YPS, Kapral MK, Fang J, Austin PC, Carbonneau A, Hill MD, Jin A, Mandzia J, Silver FL, Yu AYX. Sex Differences in Screening for Large Vessel Occlusion and Thrombectomy: A Population-Based Cohort Study. Stroke. 2026;57:2780–2788. doi: 10.1161/STROKEAHA.126.055991

2. Shah H, He T, Dyck N, Stang J, Nicol D, McIntosh CJ, Wilton SB, Coutts SB, Singh N, Hill MD, et al. Disparities in Access to Vascular Stroke Imaging and Carotid Revascularization: A Population Study. J Am Heart Assoc. 2025;14:e043715. doi: 10.1161/JAHA.125.043715

3. Grundtvig J, Sværke K, Preskou M, Christensen LM, Steiner T, Christensen H. Comparison of access to stroke diagnostics, treatment, rehabilitation, and outcome between men and women. Glob Epidemiol. 2025;9:100201. doi: 10.1016/j.gloepi.2025.100201

4. Wang JJ, Katz JM, Sanmartin M, Naidich JJ, Rula E, Sanelli PC. Gender-Based Disparity in Acute Stroke Imaging Utilization and the Impact on Treatment and Outcomes: 2012 to 2021. J Am Coll Radiol. 2024;21:128–140. doi: 10.1016/j.jacr.2023.07.015

5. Goyal M, Menon BK, van Zwam WH, Dippel DW, Mitchell PJ, Demchuk AM, Dávalos A, Majoie CB, van der Lugt A, de Miquel MA, et al. Endovascular thrombectomy after large-vessel ischaemic stroke: a meta-analysis of individual patient data from five randomised trials. Lancet. 2016;387:1723–31. doi: 10.1016/S0140-6736(16)00163-X

6. Chalos V, de Ridder IR, Lingsma HF, Brown S, van Oostenbrugge RJ, Goyal M, Campbell BCV, Muir KW, Guillemin F, Bracard S, et al. Does Sex Modify the Effect of Endovascular Treatment for Ischemic Stroke? Stroke. 2019;50:2413–2419. doi: 10.1161/STROKEAHA.118.023743

7. Xie X, Lambrinos A, Chan B, Dhalla IA, Krings T, Casaubon LK, Lum C, Sikich N, Bharatha A, Pereira VM, et al. Mechanical thrombectomy in patients with acute ischemic stroke: a cost-utility analysis. CMAJ Open. 2016;4:E316–25. doi: 10.9778/cmajo.20150088

8. Shireman TI, Wang K, Saver JL, Goyal M, Bonafé A, Diener HC, Levy EI, Pereira VM, Albers GW, Cognard C, et al. Cost-Effectiveness of Solitaire Stent Retriever Thrombectomy for Acute Ischemic Stroke: Results From the SWIFT-PRIME Trial (Solitaire With the Intention for Thrombectomy as Primary Endovascular Treatment for Acute Ischemic Stroke). Stroke. 2017;48:379–387. doi: 10.1161/STROKEAHA.116.014735

9. Sevick LK, Ghali S, Hill MD, Danthurebandara V, Lorenzetti DL, Noseworthy T, Spackman E, Clement F. Systematic Review of the Cost and Cost-Effectiveness of Rapid Endovascular Therapy for Acute Ischemic Stroke. Stroke. 2017;48:2519–2526. doi: 10.1161/STROKEAHA.117.017199

10. Heggie R, Wu O, White P, Ford GA, Wardlaw J, Brown MM, Clifton A, Muir KW. Mechanical thrombectomy in patients with acute ischemic stroke: A cost-effectiveness and value of implementation analysis. Int J Stroke. 2020;15:881–898. doi: 10.1177/1747493019879656

11. Balami JS, Ford GA, Buchan AM, Gray A, Saver JL, Candio P. Cost-effectiveness of mechanical thrombectomy for acute ischemic stroke: generalizability across implementation settings. J Stroke Cerebrovasc Dis. 2026;35:108730. doi: 10.1016/j.jstrokecerebrovasdis.2026.108730

12. Wu X, Hughes DR, Gandhi D, Matouk CC, Sheth K, Schindler J, Wira C, Wintermark M, Sanelli P, Malhotra A. CT Angiography for Triage of Patients with Acute Minor Stroke: A Cost-effectiveness Analysis. Radiology. 2020;294:580–588. doi: 10.1148/radiol.2019191238

13. Martinez G, Katz JM, Pandya A, Wang JJ, Boltyenkov A, Malhotra A, Mushlin AI, Sanelli PC. Cost-Effectiveness Study of Initial Imaging Selection in Acute Ischemic Stroke Care. J Am Coll Radiol. 2021;18:820–833. doi: 10.1016/j.jacr.2020.12.013

14. Gadsden T, Si L, Atkins ER, Carcel C, Wang X, Jan S, Woodward M, Downey LE. Health and economic benefits of improving pre-hospital identification of stroke in Australian women: a modelling study. Med J Aust. 2025;223:141–148. doi: 10.5694/mja2.52701

15. McAteer H, Cosh E, Freeman G, Pandit A, Wood P, Lilford R. Cost-effectiveness analysis at the development phase of a potential health technology: examples based on tissue engineering of bladder and urethra. J Tissue Eng Regen Med. 2007;1:343–9. doi: 10.1002/term.36

16. Husereau D, Drummond M, Augustovski F, de Bekker-Grob E, Briggs AH, Carswell C, Caulley L, Chaiyakunapruk N, Greenberg D, Loder E, et al. Consolidated Health Economic Evaluation Reporting Standards 2022 (CHEERS 2022) statement: updated reporting guidance for health economic evaluations. BMJ. 2022;376:e067975. doi: 10.1136/bmj-2021-067975

17. Goyal M, Ospel JM, Ganesh A, Dowlatshahi D, Volders D, Möhlenbruch MA, Jumaa MA, Nimjee SM, Booth TC, Buck BH, et al. Endovascular Treatment of Stroke Due to Medium-Vessel Occlusion. N Engl J Med. 2025;392:1385–1395. doi: 10.1056/NEJMoa2411668

18. Ye Z, Zhou T, Zhang M, Zhou J, Xie F, Hill MD, Smith EE, Busse JW, Zhang Y, Liu Y, et al. Cost-effectiveness of endovascular thrombectomy with alteplase versus endovascular thrombectomy alone for acute ischemic stroke secondary to large vessel occlusion. CMAJ Open. 2023;11:E443–E450. doi: 10.9778/cmajo.20220096

19. Statistics Canada. Table 13-10-0114-01. Life expectancy and other elements of the complete life table, three-year estimates, Canada, all provinces except Prince Edward Island. doi:10.25318/1310011401-eng

20. National Center for Health Statistics. United States life tables, 2024, Table 3, Life table for females. National Vital Statistics Reports 75-05. Hyattsville, MD: NCHS.

21. Rebchuk AD, O’Neill ZR, Szefer EK, Hill MD, Field TS. Health Utility Weighting of the Modified Rankin Scale: A Systematic Review and Meta-analysis. JAMA Netw Open. 2020;3:e203767. doi: 10.1001/jamanetworkopen.2020.3767

22. CADTH. Guidelines for the Economic Evaluation of Health Technologies: Canada. 4th ed. Ottawa: CADTH; 2017.

23. Sanders GD, Neumann PJ, Basu A, Brock DW, Feeny D, Krahn M, Kuntz KM, Meltzer DO, Owens DK, Prosser LA, et al. Recommendations for Conduct, Methodological Practices, and Reporting of Cost-effectiveness Analyses: Second Panel on Cost-Effectiveness in Health and Medicine. JAMA. 2016;316:1093–103. doi: 10.1001/jama.2016.12195

24. Anderson JL, Heidenreich PA, Barnett PG, Creager MA, Fonarow GC, Gibbons RJ, Halperin JL, Hlatky MA, Jacobs AK, Mark DB, et al. ACC/AHA statement on cost/value methodology in clinical practice guidelines and performance measures: a report of the American College of Cardiology/American Heart Association Task Force on Performance Measures and Task Force on Practice Guidelines. Circulation. 2014;129:2329–45. doi: 10.1161/CIR.0000000000000042

25. Centers for Medicare & Medicaid Services. Medicare Program; Hospital Inpatient Prospective Payment Systems for Acute Care Hospitals, Fiscal Year 2026 Final Rule (CMS-1833-F), Tables 1A to 1E and Table 5. Baltimore, MD: CMS; 2025.

26. Centers for Medicare & Medicaid Services. Calendar Year 2026 Physician Fee Schedule, Relative Value File RVU26D. Baltimore, MD: CMS; 2026.

27. ElHabr AK, Katz JM, Wang J, Bastani M, Martinez G, Gribko M, Hughes DR, Sanelli P. Predicting 90-day modified Rankin Scale score with discharge information in acute ischaemic stroke patients following treatment. BMJ Neurol Open. 2021;3:e000177. doi: 10.1136/bmjno-2021-000177

28. Medicare Payment Advisory Commission. Report to the Congress: Medicare Payment Policy, March 2026. Chapters 7 (skilled nursing facility services), 8 (home health care services) and 9 (inpatient rehabilitation facility services). Washington, DC: MedPAC; 2026.

29. US Bureau of Labor Statistics. Consumer Price Index for All Urban Consumers, medical care (series CUUR0000SAM), annual averages 2015 to 2025 and monthly values January to September 2026. Washington, DC: BLS.

30. Zhu J, Kamel H, Gupta A, Mushlin AI, Menzies NA, Gaziano TA, Rosenthal MB, Pandya A. Prioritizing Quality Measures in Acute Stroke Care: A Cost-Effectiveness Analysis. Ann Intern Med. 2023;176:649–657. doi: 10.7326/M22-3186

31. Hemming K, Haines TP, Chilton PJ, Girling AJ, Lilford RJ. The stepped wedge cluster randomised trial: rationale, design, analysis, and reporting. BMJ. 2015;350:h391. doi: 10.1136/bmj.h391

32. Dahabreh IJ, Robertson SE, Steingrimsson JA, Stuart EA, Hernán MA. Extending inferences from a randomized trial to a new target population. Stat Med. 2020;39:1999–2014. doi: 10.1002/sim.8426

